# Nebulized in-line endotracheal dornase alfa and albuterol administered to mechanically ventilated COVID-19 patients: A case series

**DOI:** 10.1101/2020.05.13.20087734

**Authors:** Andrew G. Weber, Alice S. Chau, Mikala Egeblad, Betsy J. Barnes, Tobias Janowitz

**Affiliations:** Division of Pulmonary, Critical Care, and Sleep Medicine, Department of Medicine, Northwell Health, 300 Community Drive, Manhasset, NY, 11030; Division of Allergy and Infectious Diseases, Department of Medicine, University of Washington and the Center for Immunity and Immunotherapies, Seattle Children’s Research Institute, 1900 9th Ave, Seattle, WA 98101; Cancer Center, Cold Spring Harbor Laboratory, 1 Bungtown Road, Cold Spring Harbor, NY 11724; Center for Autoimmune, Musculoskeletal and Hematopoietic Diseases, The Feinstein Institutes for Medical Research and the Departments of Molecular Medicine and Pediatrics, Donald and Barbara Zucker School of Medicine at Hofstra/Northwell, 350 Community Drive, Manhasset, NY, 11030; Northwell Health Cancer Institute, 450 Lakeville Road, New Hyde Park, NY 11042

**Keywords:** SARS-CoV-2, COVID-19, coronavirus, mucopurulent secretions, dornase alfa, neutrophil extracellular traps, ARDS, VV-ECMO

## Abstract

**Background:** Mechanically ventilated patients with coronavirus disease 2019 (COVID-19) have a mortality of 24-53%, in part due to distal mucopurulent secretions interfering with ventilation. Dornase alfa is recombinant human DNase 1 and digests DNA in mucoid sputum. Nebulized dornase alfa is FDA-approved for cystic fibrosis treatment. DNA from neutrophil extracellular traps (NETs) contributes to the viscosity of mucopurulent secretions. NETs are found in the serum of patients with severe COVID-19, and targeting NETs reduces mortality in animal models of acute respiratory distress syndrome (ARDS). Thus, dornase alfa may be beneficial to patients with severe COVID-19—acting as a mucolytic and targeting NETs. However, delivery of nebulized drugs can aerosolize SARS-CoV-2, which causes COVID-19, increasing the infection risk for staff. Here, we report a single center case series where dornase alfa was administered through an in-line nebulizer system to minimize risk of virus aerosolization.

**Methods:** Demographic, clinical data, and outcomes were collected from the electronic medical records of five mechanically ventilated patients with COVID-19—including three requiring veno-venous extracorporeal membrane oxygenation (VV-ECMO)—treated with nebulized in-line endotracheal dornase alfa co-administered with albuterol (used to increase delivery to the alveoli), between March 31 and April 24, 2020. Data on tolerability and responses, including longitudinal values capturing respiratory function and inflammatory status, were analyzed.

**Results:** Following nebulized in-line administration of dornase alfa with albuterol, the fraction of inspired oxygen requirements was reduced for all five patients. All patients remain alive and two patients have been discharged from the intensive care unit. No drug associated toxicities were identified.

**Conclusions:** The results presented in this case series suggest that dornase alfa will be well-tolerated by critically ill patients with COVID-19. Clinical trials are required to formally test the dosing, safety, and efficacy of dornase alfa in COVID-19, and two have recently been registered (*NCT04359654* and *NCT04355364*). With this case series, we hope to contribute to the development of management approaches for critically ill patients with COVID-19.

## BACKGROUND

Critically ill patients with coronavirus disease 2019 (COVID-19), caused by the severe acute respiratory syndrome coronavirus 2 (SARS-CoV-2), progress to hypoxemic and then mixed respiratory failure, secondary to acute respiratory distress syndrome (ARDS) (1, 2). Approximately 79–88% of patients admitted to the intensive care unit (ICU) with COVID-19 require intubation and mechanical ventilation, with a mortality of 24–53% (3-6). ARDS in COVID-19 is characterized by ventilation failure, in part attributable to distally located mucopurulent secretions.

Dornase alfa (Pulmozyme®) is recombinant human DNase 1 and a safe mucolytic that is administered in nebulized form. It is FDA-approved in combination with standard therapies for patients with cystic fibrosis to improve sputum clearance and pulmonary function (7). It is also used off-label as a mucolytic in other diseases, including ARDS (8, 9). A mechanism by which dornase alfa might improve ventilation is by reducing the DNA-mediated viscosity of neutrophil-rich secretions (10). There are multiple sources for the DNA in mucoid sputum, one of which is neutrophil extracellular traps (NETs). Recently, we collaboratively reported that in the discarded serum of patients with COVID-19, the levels of NETs were increased and were correlated with lactate dehydrogenase (LDH), D-dimer, and C-reactive protein (CRP) levels (11). Targeting NETs reduces mortality in animal models of ARDS (12). Despite recognition that mucolytic treatment may be beneficial for patients with COVID-19, administration of nebulized medications, such as dornase alfa, have been limited due to risk of viral aerosolization. If risk of viral aerosolization can be avoided, dornase alfa may benefit patients with severe COVID-19. by acting as a mucolytic and by reducing NET levels in the lungs, thereby improving oxygenation and ventilation. We report the clinical course, safety, and outcomes after nebulized in-line endotracheal dornase alfa treatment for five intubated and mechanically ventilated patients with PCR-confirmed COVID-19.

## METHODS

The Northwell Health institutional review board that focuses on COVID-19 research approved this case series as minimal-risk research using de-identified data from routine clinical practice. Data were collected from the enterprise health record (Sunrise Clinical Manager; Allscripts) reporting database, and included patient demographics, comorbidities, inpatient medications, laboratory studies, treatment, and outcomes. We further obtained longitudinal values of FiO_2_ and of the arterial partial pressure of carbon dioxide (PaCO_2_) as measures of respiratory function during treatment. FiO_2_ values of the circuit were reported for those patients who required veno-venous extracorporeal membrane oxygenation (VV-ECMO). Ferritin, CRP, LDH, and D-dimer were obtained as measures of systemic disease and inflammation. Not all patients had laboratory investigations on the same days in relation to the nDA+A treatment. In the following case synopses, each measurement is therefore followed by the day in relation to the first day of treatment with nDA+A (*e.g*. d 2 for the second day of treatment with nDA+A or d -1 for the day before nDA+A treatment was initiated).

## RESULTS

Five patients treated with dornase alfa between March 31, 2020 and April 24, 2020 were identified. These patients had met the Berlin criteria for ARDS and were treated with ventilator strategies guided by the ARDSNet protocol at North Shore University Hospital within Northwell Health (13). They had been treated with dornase alfa because they required high levels of fraction of inspired oxygen (FiO_2_) and had elevated ventilation demands. All patients received the same treatment doses: nebulized dornase alfa (2.5 mg) co-administered twice daily with the short-acting β_2_-agonist albuterol (2.5 mg, hereafter abbreviated as nDA+A) to improve delivery to the alveoli. Of note, β_2_-adrenoreceptor agonism may also inhibit NET formation by direct action on neutrophils (14). The treatment was administered with an Aerogen® Solo in-line nebulizer to avoid open aerosol generation, which would place staff at risk of exposure to SARS-CoV-2.

The patient characteristics are summarized in **Table 1**. Patients were treated with nDA+A between 3 to 25 days. The most common characteristics of the patients included obesity (BMI≥30) and four of the patients had hypertension. Four patients received methylprednisolone dosed at 1-2mg/kg/day. All patients were treated with full dose or prophylactic dose anticoagulation for thrombosis. All other medications that were administered during the course of hospitalization are summarized in **Table S1**. The clinical course of the five patients treated with nDA+A is summarized in **Figure 1. Figure 2** and **3** display the longitudinal, ventilatory, and inflammatory markers for each patient.

**Table 1.**
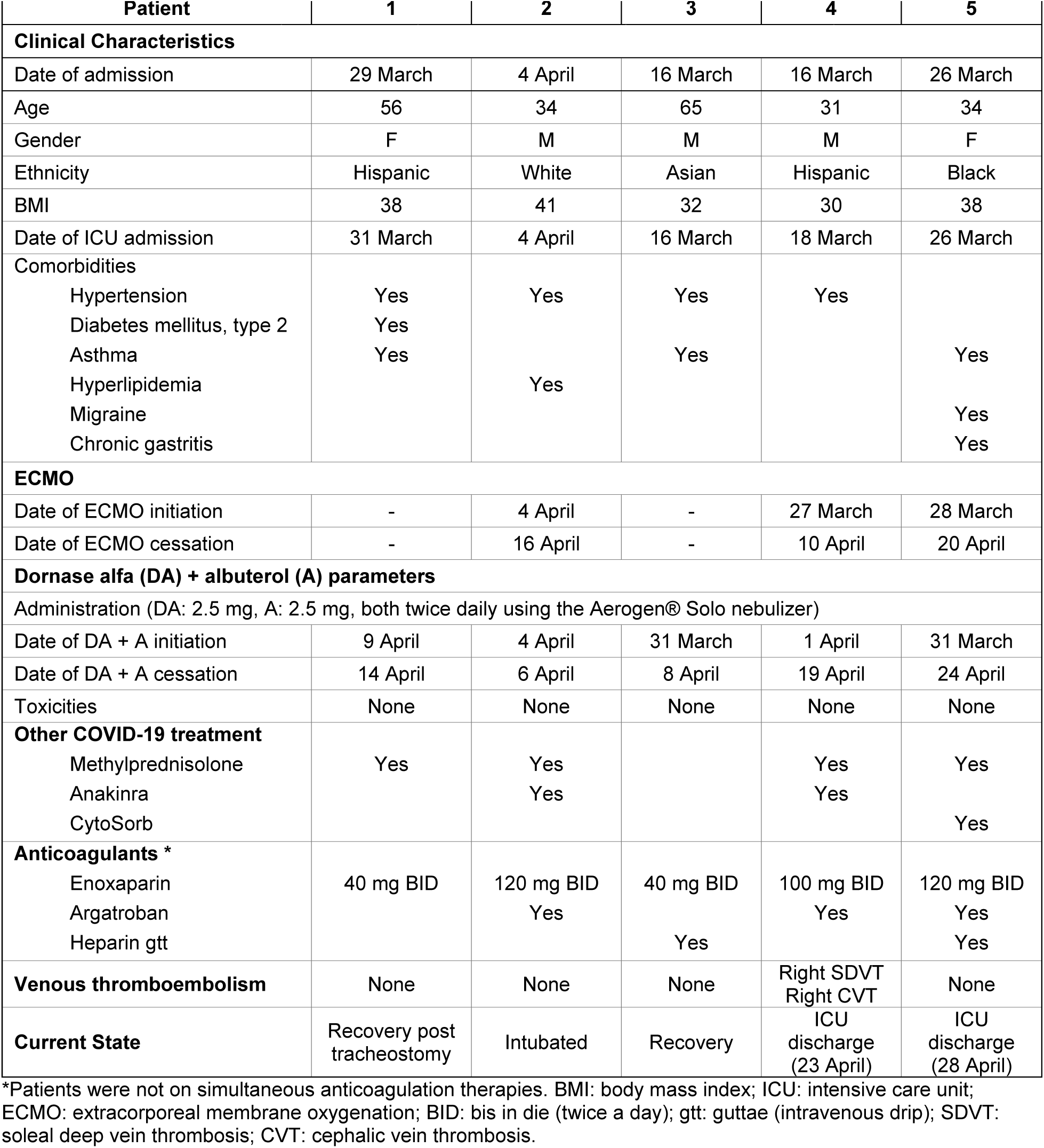
Patient data from five patients with COVID-19 who received dornase alfa with albuterol March-April, 2020.

**Figure 1.**
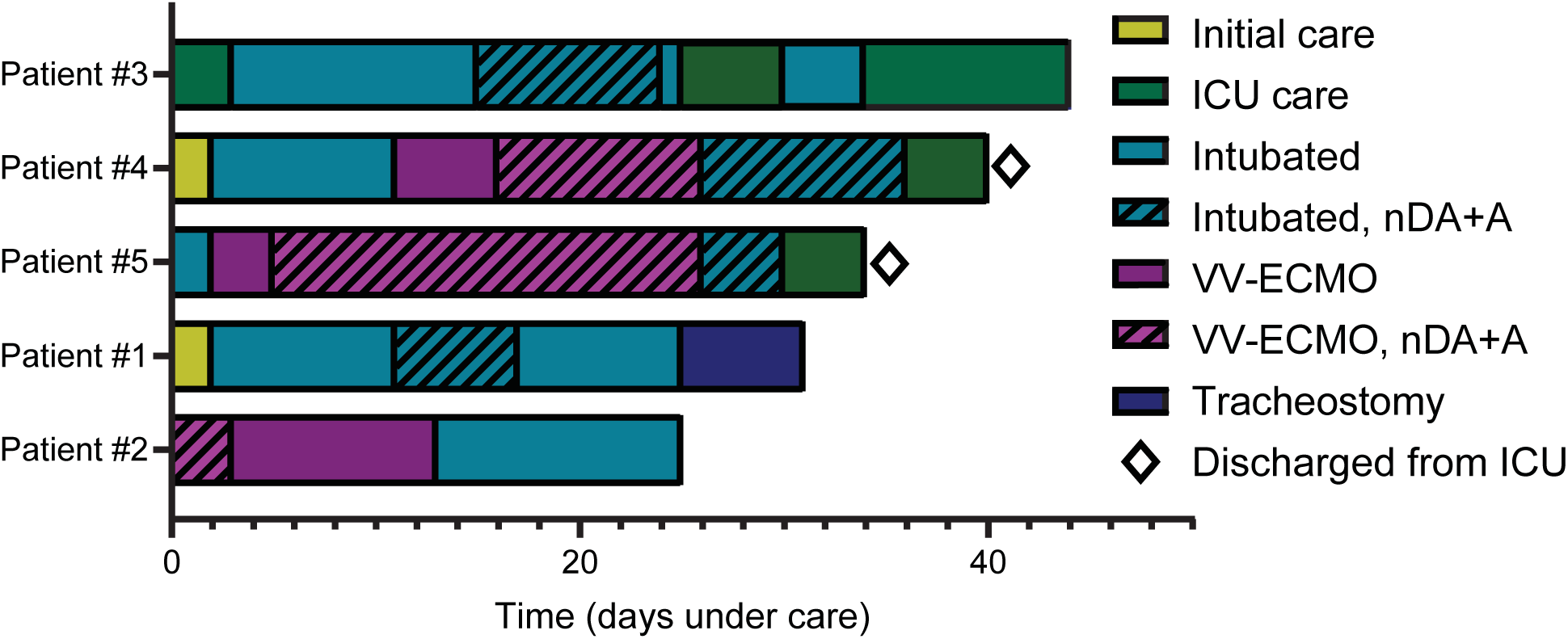
Overview of the clinical course of five patients treated with nebulized dornase alfa + albuterol (nDA+A).

**Figure 2.**
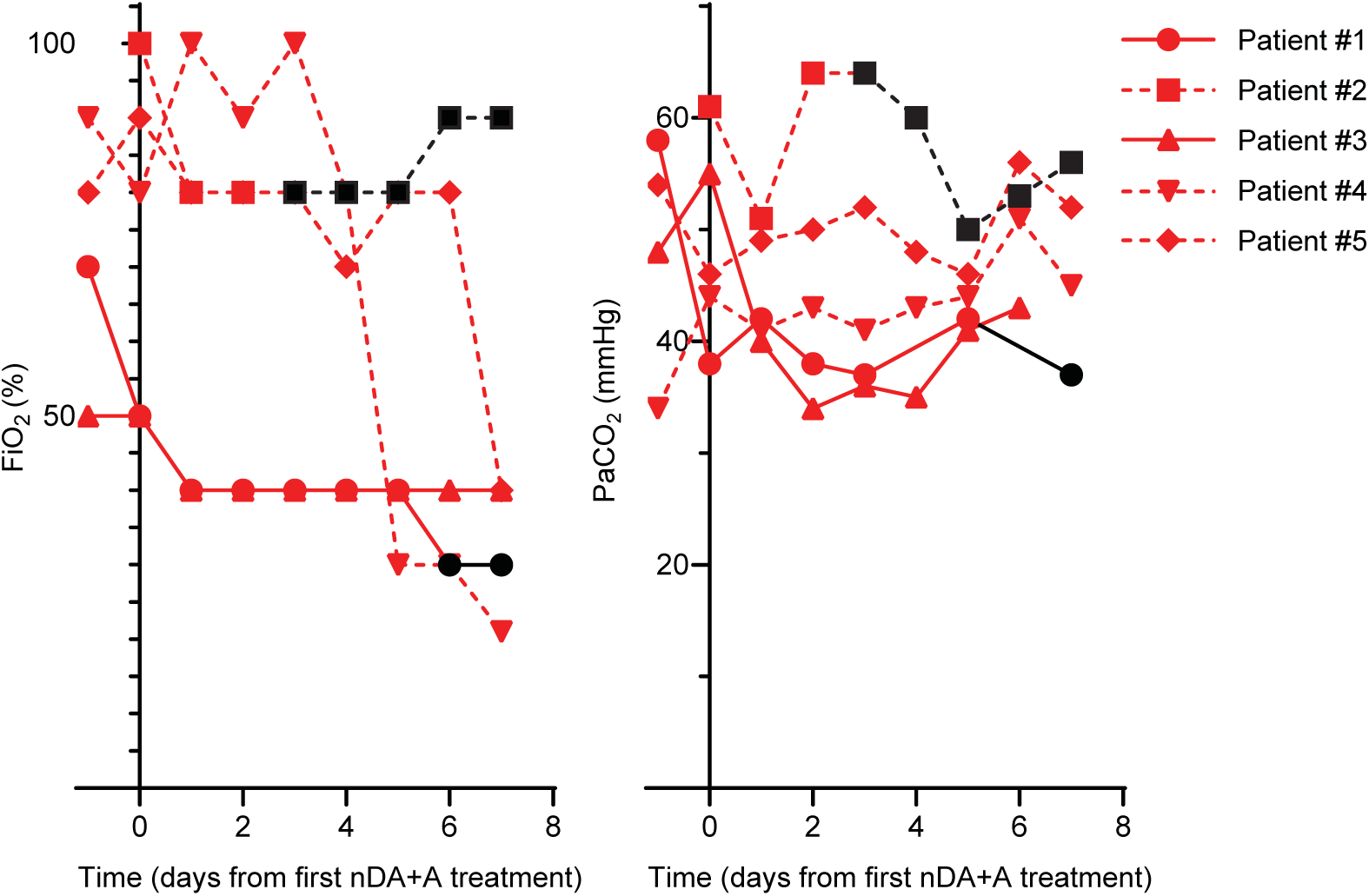
Patient-level data of respiratory function during treatment with nebulized dornase alfa + albuterol (nDA+A). Values were extracted from the medical records the day before and up to the seven days after the initiation of treatment. Values are graphed in black for patients after they ceased nDA+A treatment. Dashed lines indicate patients on VV-ECMO. Not all markers were measured daily for every patient. FiO_2_: fraction of inspired oxygen; PaCO_2_: partial pressure of carbon dioxide.

**Figure 3.**
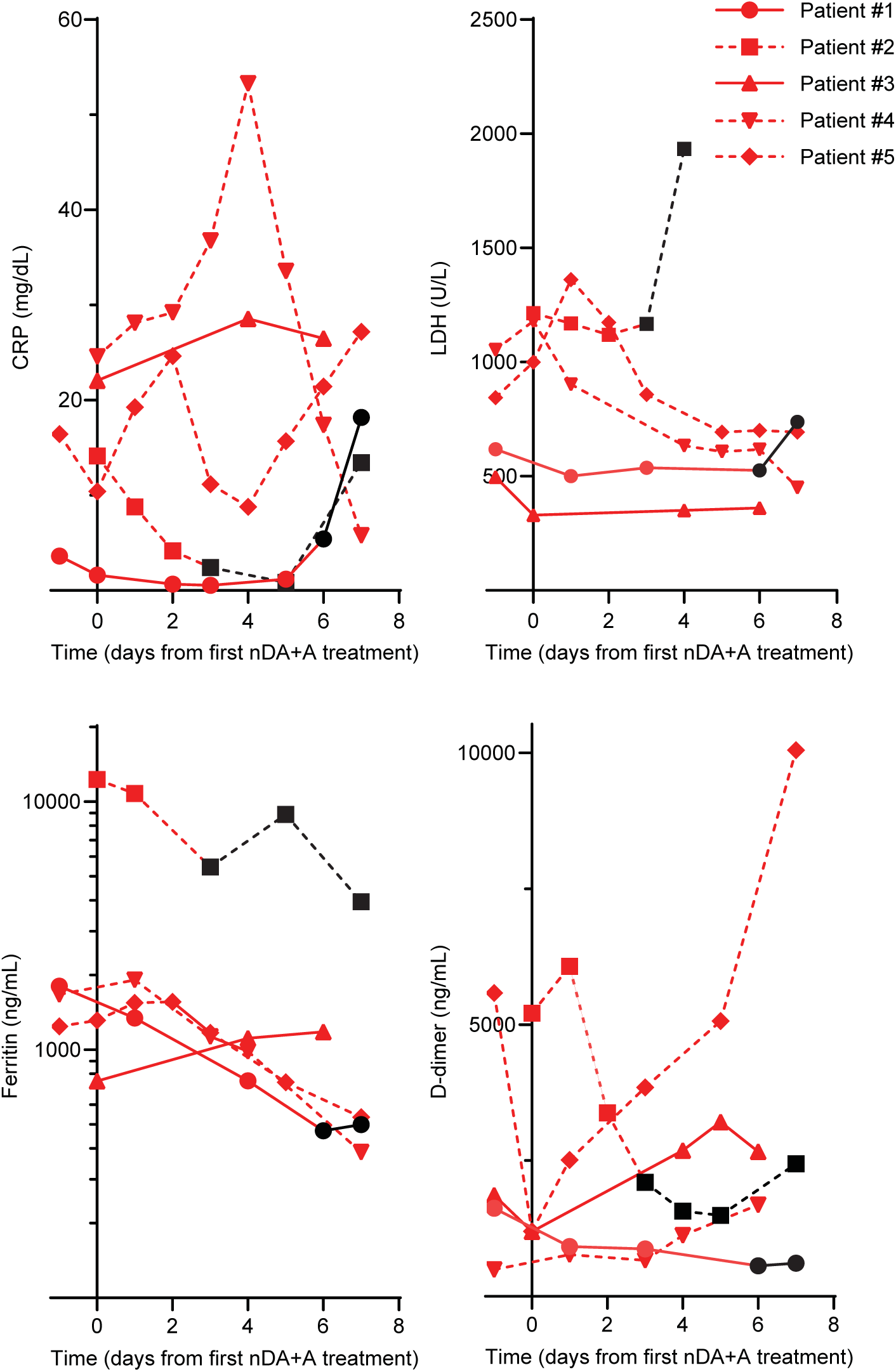
Patient-level data of systemic disease during treatment with nebulized dornase alfa + albuterol (nDA+A). Values were extracted from the medical records the day before and up to the seven days after the initiation of treatment. Values are graphed in black for patients after they ceased nDA+A treatment. Dashed lines indicate patients on VV-ECMO. Not all markers were measured daily for every patient. CRP: C-reactive protein; LDH: lactate dehydrogenase.

Patient 1 is a 56-year-old Hispanic woman who presented in respiratory distress. Her respiratory status deteriorated over 48 hours, requiring intubation and transfer to the ICU. She was treated with nDA+A for six days, starting from day 9 of intubation. The FiO_2_ requirement decreased from 70% (d -1) to 30% (d 6), PaCO_2_ from 58 (d -1) to 37 mmHg (d 7), ferritin from 1,803 (d -1) to 472 ng/mL (d 6), and D-dimer from 1,619 (d -1) to 563 ng/mL (d 6). Minimal changes were noted in CRP and LDH. The patient underwent a tracheostomy after 23 days of endotracheal intubation and remains on an FiO_2_ of 30% while pending return of mental status.

Patient 2 is a 34-year-old white man who presented to the hospital in diabetic ketoacidosis without prior history of diabetes mellitus. He was intubated on admission and initiated on VV-ECMO. He received nDA+A for three days and was de-cannulated after 12 days. The FiO_2_ requirement decreased from 100% (d 0) to 80% (d 3), CRP from 14.14 (d 0) to 2.41 mg/dL (d 3), ferritin from 12,281 (d 0) to 5,453 ng/mL (d 3), and D-dimer from 5,210 (d 0) to 2,099 ng/mL (d 3). Minimal changes were noted in PaCO_2_ and LDH. The patient remains intubated.

Patient 3 is a 65-year-old Asian man who was admitted directly to the ICU for respiratory distress and intubated three days later. Twelve days after intubation, he was started on nine days of nDA+A treatment. The FiO_2_ requirement decreased from 50% (d -1) to 40% (d 7), PaCO_2_ from 55 (d 0) to 43 mmHg (d 6), and CRP from 22.07 (d 0) to 26.48 mg/dL (d 6). Minimal changes were noted in ferritin, LDH, and D-dimer. He was extubated one day after the completion of the nDA+A course. Six days later, he was re-intubated for an additional four days due to mental status changes and failure to protect his airway. The patient remains extubated in ICU care.

Patient 4 is a 31-year-old Hispanic man who was intubated and transferred to the ICU from the Internal Medicine service two days after presenting with respiratory distress. Nine days after intubation, he was initiated on VV-ECMO. Five days after cannulation, he was started on the nDA+A treatment. After nine days, he was de-cannulated and remained intubated for ten days while continuing the nDA+A treatment. He was then extubated and discharged to the floor. The FiO_2_ requirement decreased from 90% (d -1) to 21% (d 7) and LDH from 1,054 (d -1) to 451 U/L (d 7). Ferritin initially decreased from 1,669 (d -1) to 387 ng/mL (d 7). On day 15 of treatment, he developed methicillin-resistant *Staphylococcus aureus* (MRSA) pneumonia and bacteremia. Ferritin thus increased to 1,619 ng/mL (d 13) prior to decreasing to 555 ng/mL (d 19) with antibiotic treatment. Minimal changes were noted in PaCO_2_, CRP, and D-dimer.

Patient 5 is a 34-year-old black woman who was intubated at an outside hospital, then transferred to the North Shore University Hospital ICU. Two days later, she was cannulated for VV-ECMO. She required VV-ECMO for 13 days and was intubated for a total of 29 days. She was treated with nDA+A for 25 days starting three days following intubation and cannulation. While on VV-ECMO for the first five days, CytoSorb therapy was applied. She was de-cannulated after 23 days, extubated after 4 days, and discharged to the floor. The FiO_2_ requirement fell from 80% (d -1) to 40% (d 7), ferritin from 1,244 (d -1) to 535 ng/mL (d 7), and LDH from 844 (d -1) to 693 U/L (d 7). Minimal changes were noted in PaCO_2_, CRP, and D-dimer.

## DISCUSSION

At the doses utilized, no nDA+A treatment-associated toxicities were identified. FiO_2_ requirements decreased for all five patients seven days after nDA+A treatment was initiated. All patients remain alive at the time of submission of this report, with two patients discharged from the ICU. We recognize that these FiO_2_ changes may be independent of the nDA+A treatment. Clinical trials are therefore required to test the dose range, safety, and efficacy of dornase alfa in patients with COVID-19 in this setting and possibly earlier in the disease course. Endpoints should include measurements of the effect on respiratory function as well as on systemic inflammation, coagulopathy, secondary infections, and the presence of NETs in plasma. Two such trials were recently registered (*NCT04359654* and *NCT04355364*).

It is not clear whether nebulized dornase alfa will have any effect on blood NET levels or systemic inflammation in COVID-19, but a reduction in systemic inflammatory markers has been reported after use of dornase alfa in patients with cystic fibrosis (7). We did note a reduction in CRP in two patients (patients 2 and 3) and a reduction in D-dimer in two patients (patients 1 and 2) during nDA+A treatment. LDH was reduced for the patients on VV-ECMO during nDA+A treatment, and ferritin was reduced in four out of five patients. Due to the small sample size and the common occurrence of secondary infections in ventilated patients with COVID-19, we are unable to comment on any potential relationship between nDA+A administration and the risk of secondary infections.

## CONCLUSIONS

Nebulized dornase alfa in combination with albuterol may be a safe treatment option for mechanically ventilated patients with ARDS secondary to COVID-19, including for those on VV-ECMO—a patient population with an urgent, unmet need for effective therapies.

## Data Availability

All data generated or analyzed during this study are included within the article.

## LIST OF ABBREVIATIONS

ARDS: acute respiratory distress syndrome
BID: bis in die (twice daily)
COVID-19: coronavirus disease 2019
CRP: C-reactive protein
d: day
FiO_2_: fraction of inspired oxygen
gtt: guttae (intravenous drip)
ICU: intensive care unit
LDH: lactate dehydrogenase
MRSA: methicillin-resistant *Staphylococcus aureus*
nDA+A: nebulized dornase alfa plus albuterol
NETs: neutrophil extracellular traps
PaCO_2_: arterial partial pressure of carbon dioxide
SARS-CoV-19: severe acute respiratory syndrome coronavirus 2
VV-ECMO: veno-venous extracorporeal membrane oxygenation

## DECLARATIONS

### Ethics approval and consent to participate

The Northwell Health institutional review board that focuses on COVID-19 research approved this case series as minimal-risk research using de-identified data from routine clinical practice. Informed consent to participate in the study was obtained from the participants or their health care proxies. The study has been registered as “Dornase Alfa Administered to Patients With COVID-19 (DACOVID)” at ClinicalTrials.gov with ClinicalTrials.gov Identifier: NCT04387786.

### Consent for publication

Not applicable.

### Availability of supporting data

All data generated or analyzed during this study are included within the article.

### Competing interests

Mikala Egeblad is receiving lonodelestat from Santhera for preclinical studies, but has no financial relationship with Santhera. The other authors declare that they have no competing interests.

### Funding

This work was supported by the William C. and Joyce C. O’Neil Charitable Trust. In addition, M.E. and T.J. are supported by NIH grant 5P30CA045508-30. B.J.B. is supported by NIH grant 1R01AR076242-01 and DOD LRP W81XWH-18-1-0674. A.S.C. was supported by The Primary Immune Deficiency Treatment Consortium (U54 AI 082973), funded jointly by NCATS (write out) and the National Institute of Allergy and Infectious Diseases (NIAID). The funding bodies had no role in the design of the study; in collection, analysis, and interpretation of data; and in writing the manuscript.

### Authors’ contributions

*Concept and design, analysis and interpretation of data, and drafting of the manuscript:* A.G.W., A.S.C., M.E., B.J.B., and T.J. *Data acquisition:* A.G.W. and B.J.B.

## Acknowledgements

The authors thanks “The NETwork to Target Neutrophils in COVID-19,” Eric Gottesman, William Taylor, David Menon, and David Tuveson for helpful discussions, as well as the Northwell COVID-19 Research Consortium for facilitating the study.

## Authors’ information

Not applicable.

## Disclaimer

The initial characteristics of 5,700 patients from Northwell Health are presented elsewhere (5). This case series presented in-depth results on the clinical status of five patients treated with dornase alfa that were not presented in that article.

---

**Supplemental Table 1.** Additional medications that dornase alfa+albuterol-treated COVID-19 patients received while in the hospital.

| Patient | 1 | 2 | 3 | 4 | 5 |
| --- | --- | --- | --- | --- | --- |
| **Hospital medications** |  |  |  |  |  |
| Amiodarone |  | Yes |  | Yes |  |
| Ampicillin |  |  |  |  | Yes |
| Ascorbic acid |  |  | Yes | Yes | Yes |
| Azithromycin | Yes |  | Yes | Yes |  |
| Bumetanide |  | Yes | Yes |  |  |
| Caspofungin |  |  | Yes | Yes | Yes |
| Cefepime | Yes |  |  | Yes | Yes |
| Ceftriaxone |  |  |  | Yes |  |
| Cisatracurium |  |  | Yes | Yes | Yes |
| Dexmedetomidine | Yes |  | Yes | Yes | Yes |
| Dobutamine | Yes |  |  |  |  |
| Esmolol |  |  |  | Yes |  |
| Fentanyl | Yes | Yes | Yes | Yes | Yes |
| Fluconazole | Yes |  | Yes |  |  |
| Fosphenytoin | Yes |  |  |  |  |
| Furosemide | Yes |  | Yes | Yes | Yes |
| HCQ/CQ | Yes |  | Yes | Yes | Yes |
| Hydromorphone |  |  |  | Yes |  |
| Insulin | Yes | Yes | Yes | Yes | Yes |
| IVIG |  | Yes |  |  |  |
| Ketamine | Yes | Yes | Yes | Yes | Yes |
| Levetiracetam | Yes |  |  |  |  |
| Meropenem |  | Yes | Yes | Yes | Yes |
| Metronidazole |  | Yes |  | Yes |  |
| Midazolam | Yes | Yes | Yes | Yes | Yes |
| Milrinone |  | Yes |  |  |  |
| Nicardipine |  |  |  | Yes |  |
| Nitroprusside |  |  |  | Yes |  |
| Norepinephrine | Yes | Yes | Yes | Yes | Yes |
| Pantoprazole | Yes | Yes | Yes | Yes | Yes |
| Phenylephrine | Yes | Yes | Yes | Yes |  |
| Propofol | Yes | Yes | Yes | Yes | Yes |
| Rocuronium | Yes | Yes | Yes |  | Yes |
| Sodium bicarbonate |  |  |  | Yes |  |
| TPN |  |  | Yes |  |  |
| Vancomycin |  | Yes | Yes | Yes | Yes |
| Vasopressin |  | Yes |  |  | Yes |
| Vecuronium |  |  |  | Yes | Yes |
| Zosyn |  |  | Yes | Yes |  |

Note: All antimicrobials were given at treatment doses. HCQ: hydroxychloroquine; CQ: chloroquine; IVIG: intravenous immunoglobulin; TPN: total parenteral nutrition

## Notes

### Clinical Trial

ClinicalTrials.gov Identifer: NCT04387786
The study is a retrospective case study.

### Funding Statement

This work was supported by the William C. and Joyce C. ONeil Charitable Trust. In addition, M.E. and T.J. are supported by NIH grant 5P30CA045508-30. B.J.B. is supported by NIH grant 1R01AR076242-01 and DOD LRP W81XWH-18-1-0674. A.S.C. was supported by The Primary Immune Deficiency Treatment Consortium (U54 AI 082973), funded jointly by NCATS (write out) and the National Institute of Allergy and Infectious Diseases (NIAID).

